# Sexual risk behaviours following medical male circumcision: a matched pseudo-cohort analysis using population-based survey data

**DOI:** 10.64898/2026.04.11.26350676

**Authors:** David Mwakazanga, Victor Daka, Johnathan Gwasupika, Aliness Dombola, Kelvin Kapungu, Shepherd Khondowe, Gershom Chongwe, Isaac Fwemba, Emmanuel Ogundimu

**Affiliations:** Department of Public Health, National Health Research and Training Institute, P. O. Box 71769, Ndola, Zambia; Department of Public Health, Michael Chilufya Sata School of Medicine, The Copperbelt University, P. O. Box 71191, Ndola. Zambia; Department of Public Health, School of Medicine, The University of Zambia, P. O. Box 50110, Lusaka. Zambia; Department of Mathematical Sciences, Durham University, Stockton Road Durham, DH1 3LE, England

## Abstract

Medical male circumcision (MMC) is an established HIV prevention intervention, yet concerns persist that circumcised men may adopt higher-risk sexual behaviours following the procedure. Evidence from observational studies has been inconsistent, partly because many analyses do not adequately distinguish behaviours that occur before circumcision from those that occur afterward. This study assessed the association between MMC and subsequent sexual behaviours while demonstrating how population-based cross-sectional survey data can be adapted to address this temporal challenge. We analysed nationally representative data from the 2024 Zambia Demographic and Health Survey (ZDHS), including men aged 15–59 years who reported their circumcision status. Men who had undergone medical circumcision were compared with uncircumcised men using a matched pseudo-cohort framework that reconstructed temporal ordering based on age at circumcision. Propensity score overlap weighting was applied to improve comparability between circumcised and uncircumcised men, and odds ratios were estimated using logistic regression models incorporating overlap weights and accounting for the complex survey design. Sexual behaviour outcomes occurring after circumcision included condom non-use at last sexual intercourse, multiple sexual partners in the past 12 months, self-reported sexually transmitted infection (STI) symptoms, and composite measures of sexual risk behaviour. The analysis included 9,609 men, of whom 33.3% were medically circumcised. MMC was associated with lower odds of condom non-use at last sexual intercourse (adjusted odds ratio [aOR] = 0.75, 95% confidence interval [CI]: 0.67–0.85) and lower odds of reporting any sexual risk behaviour (aOR = 0.83, 95% CI: 0.72–0.95). No meaningful associations were observed between MMC and reporting multiple sexual partners, self-reported STI symptoms, or higher levels of composite sexual risk behaviour. In this population-based study, MMC was not associated with sexual risk compensation under routine programme conditions within the overlap population defined by the weighting scheme, supporting the behavioural safety of MMC and illustrating the value of explicitly addressing temporality when analysing behavioural outcomes using cross-sectional survey data.

## INTRODUCTION

Voluntary medical male circumcision (VMMC) has been a key component of HIV combination prevention strategies in sub-Saharan Africa since the World Health Organisation (WHO) and the Joint United Nations Programme on HIV/AIDS (UNAIDS) endorsed it in the mid-2000s based on evidence that it reduces female-to-male HIV acquisition by approximately 60 % (1,2). WHO guidance continues to recommend VMMC as an efficacious addition to comprehensive HIV prevention programming for adolescent boys and adult men in high-prevalence settings (2).

Although the biological efficacy of medical male circumcision (MMC) for HIV prevention is well established, concerns have persisted that circumcised men may perceive themselves as partially protected against HIV and consequently engage in higher-risk sexual practices, a phenomenon commonly described as behavioural risk compensation (3). Evidence from observational studies has been mixed. While several population-based and cohort analyses report no meaningful increases in condomless sex or multiple partnerships after circumcision (3–6), others suggest modest to significant risky sexual behaviour patterns among circumcised men (7–9). This heterogeneity has sustained debate about whether conclusions drawn from early randomised trials remain applicable to contemporary, large-scale circumcision programmes operating under routine health-system conditions.

A key limitation underlying these mixed findings lies in the analytic treatment of sexual behaviour within observational study designs, rather than in the data sources themselves. In many observational analyses of MMC and HIV-related outcomes—including those drawing on population-based surveys—sexual behaviours measured at the time of data collection are included as covariates in regression models evaluating circumcision and HIV risk (10–12). Such approaches implicitly treat behaviours measured at survey as baseline determinants, assuming that they precede circumcision and independently influence both circumcision uptake and subsequent outcomes. (13,14). From a causal inference perspective, however, many of these behaviours may occur after circumcision and therefore represent post-exposure outcomes or mediating pathways rather than confounders. Conditioning on such variables can introduce post-treatment bias, distort effect estimates, and contribute to apparent inconsistencies between observational findings and evidence from randomised and prospective cohort studies (15,16).

The Demographic Health Survey (DHS) provide nationally representative data on men’s circumcision status and sexual behaviours, and in some settings include HIV biomarker results, which can be used to examine post-circumcision risk behaviours at scale (17,18). When age at medical circumcision is available, it is possible to construct a matched pseudo-cohort that aligns follow-up time between circumcised and uncircumcised men, enabling more plausible temporal ordering of exposure and behavioural outcomes. This approach aligns with efforts in causal inference to emulate target trials using observational data by clearly defining baseline, exposure timing, and follow-up (19).

Despite the potential of such designs, few studies have employed matched pseudo-cohorts with survey weighting and biomarker linkage to investigate behavioural outcomes post-circumcision. In this study, we constructed a matched pseudo-cohort from DHS data to evaluate post-circumcision sexual behaviour patterns among medically circumcised men. Although HIV biomarker data are available in DHS, the present analysis focused exclusively on behavioural outcomes. By explicitly anchoring baseline at age of circumcision, aligning follow-up time across exposure groups, and treating behaviours as outcomes, we aim to provide methodologically robust evidence on whether MMC is associated with sexual risk compensation under real-world programme conditions.

## METHODS

### Study design and data source

This study used data from the DHS conducted in Zambia in 2024, a nationally representative survey. The DHS employs a stratified two-stage cluster sampling design and collects detailed information on male circumcision, sexual behaviour, health service use, and sociodemographic characteristics. Although the DHS is cross-sectional, the availability of age at medical circumcision allowed us to construct a cohort-like analytic framework that preserves temporal ordering between circumcision and subsequent sexual behaviours. The data used in this study were accessed for research purposes on 24/11/2025.

### Study population

The analysis included men aged 15–59 years who completed the male questionnaire and reported their circumcision status. Men who reported traditional circumcision were excluded from the exposed group because traditional procedures vary widely in timing, technique, and associated counselling, and therefore differ fundamentally from standardized medical male circumcision services. Men with missing information on circumcision status or baseline age were excluded during pseudo-cohort construction.

### Exposure definition: medical male circumcision

The exposure of interest was MMC, defined as circumcision performed by a trained medical provider in a formal health-care setting. Men who reported having undergone medical circumcision were classified as circumcised, while men who reported no circumcision were classified as uncircumcised. Age at medical circumcision was recorded only for medically circumcised men and served as the temporal anchor for baseline definition.

### Baseline definition and pseudo-cohort construction

#### Baseline age

To establish a meaningful baseline in the absence of longitudinal follow-up, baseline age was defined as the age at which exposure occurred. Among medically circumcised men, baseline age corresponded to the reported age at circumcision. Uncircumcised men do not have a natural exposure age; therefore, baseline age for these men was assigned through matching.

This approach reflects the counterfactual comparison of circumcised men with similar men who were eligible for circumcision at the same age but remained uncircumcised. This assumption is consistent with causal inference approaches that emulate hypothetical treatment assignment in observational data (20).

#### Matching strategy

A matched pseudo-cohort was constructed by selecting two uncircumcised men for each circumcised man, matched exactly on current age at the time of survey interview. Matching was performed within single-year age strata and did not involve baseline age or any post-exposure variables. Exact matching on current age ensures that circumcised and uncircumcised men had comparable calendar-time opportunities for exposure and behavioural observation, thereby aligning follow-up periods across exposure groups and reducing the potential for immortal time bias in the reconstructed pseudo-cohort design (21). Within each age stratum, circumcised and uncircumcised men were randomly ordered, and matching was carried out without replacement.

Under this design, uncircumcised men matched to circumcised men represent individuals who had the same age at interview and comparable opportunities to undergo circumcision at the assigned baseline age but did not receive the procedure. This matching approach therefore approximates the comparison that would arise in a randomised trial in which circumcision could be assigned at a specific age.

For each circumcised man who could be matched, his baseline age was assigned to the two matched uncircumcised men. Circumcised men for whom two eligible uncircumcised men of the same age were not available were excluded. In total, 425 medically circumcised men (11.3%) were excluded at this stage due to lack of eligible matches. An additional 149 medically circumcised men (4.0%) were excluded because of a time less than 1 year since circumcision.

This process yielded a 1:2 matched pseudo-cohort in which follow-up time progressed from a shared baseline anchor. Ratios greater than one increase precision by incorporating additional control information, while gains in efficiency diminish beyond approximately three to four controls per exposed individual (22–24). The resulting sample selection process is summarised in **Fig 1**.

**Fig 1.**
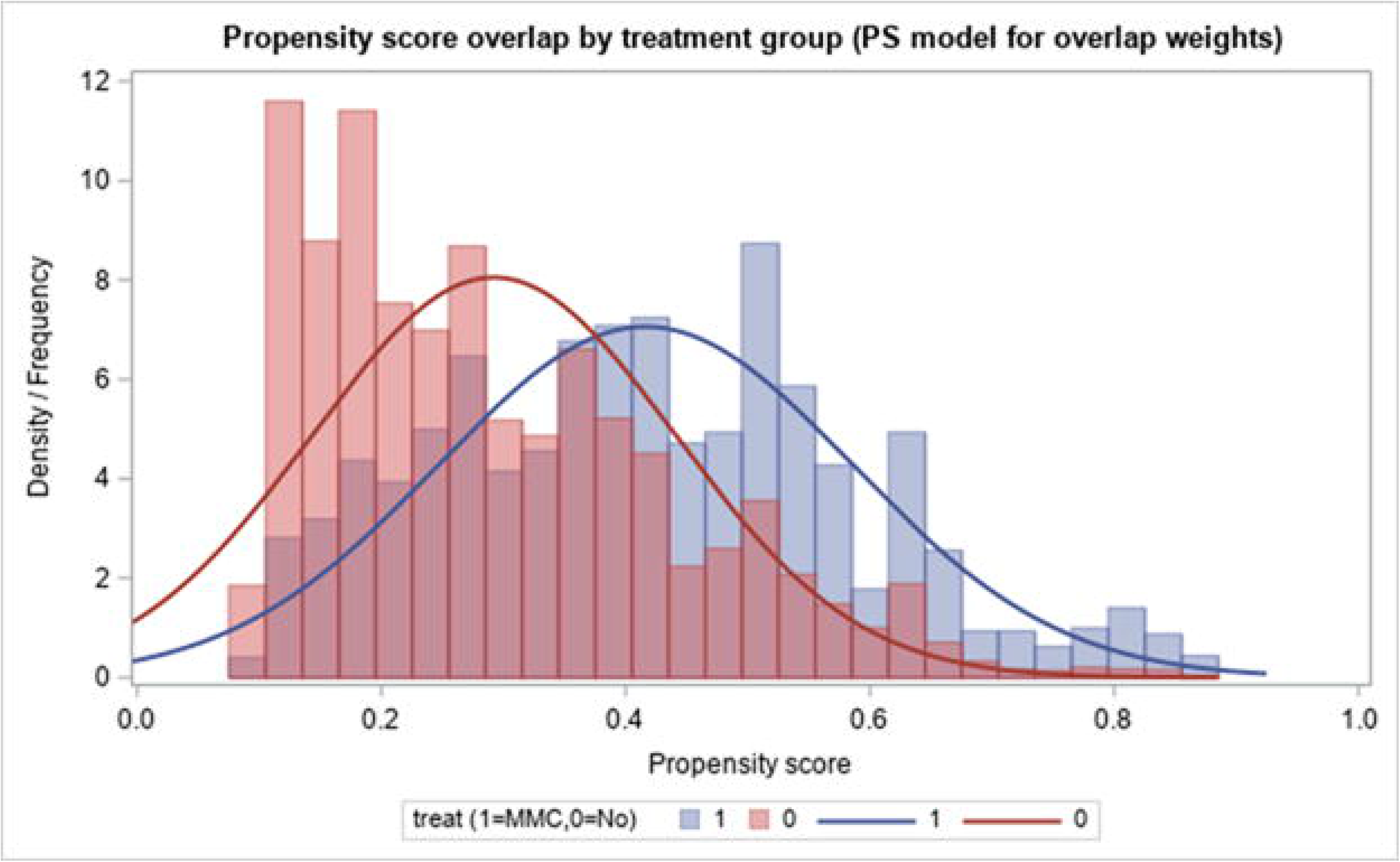
Sample selection flow diagram for the matched pseudo-cohort derived from the Zambia Demographic and Health Survey. The diagram shows the sequential steps used to construct the analytic sample from all male respondents aged 15–59 years. Men reporting traditional circumcision or missing circumcision status were excluded. Medically circumcised men were matched to uncircumcised men in a 1:2 ratio based on age at interview. The final matched pseudo-cohort was used for subsequent analyses.

To assess potential selection bias arising from exclusion of medically circumcised men without eligible age-matched controls, we compared characteristics of circumcised men retained in the analytic cohort with those excluded during pseudo-cohort construction (**Table S1**). As expected, excluded circumcised men were younger and had shorter potential follow-up time since circumcision, whereas the distributions of region, residence, religion and household wealth were broadly similar between groups. These differences reflect the temporal alignment required by the matched pseudo-cohort design, which necessitates comparable baseline age and sufficient follow-up time across exposure groups. As such, the observed differences arise from the structural requirements of the analytic framework rather than systematic socioeconomic or geographic selection, indicating that the exclusions are unlikely to meaningfully bias the estimated associations.

### Follow-up time

The primary outcomes were sexual behaviours hypothesised to occur after circumcision and potentially reflect behavioural adaptation following the procedure. These included condom use at last sexual intercourse, number of sexual partners in the past 12 months, and self-reported symptoms of sexually transmitted infections (STIs) in the past 12 months. The DHS STI indicator is based on self-reported symptoms during the preceding 12 months, including genital sores or ulcers and genital discharge. Because these measures rely on symptom recognition and self-report rather than laboratory confirmation, they may be affected by recall error, under-reporting, and misclassification and should therefore be interpreted as proxies for symptomatic STIs rather than clinically confirmed diagnoses (25). Each outcome was analysed separately to capture distinct dimensions of sexual risk behaviour.

Because the DHS collects behavioural information at the time of survey using defined recall periods, these outcomes refer to recent behaviours preceding the interview. Partner number and STI symptoms relate to the previous 12 months, whereas condom use refers to the most recent sexual intercourse. In the reconstructed pseudo-cohort, baseline corresponded to age at circumcision for circumcised men and the assigned baseline age for matched uncircumcised men. Men with less than one year of follow-up after circumcision were excluded, ensuring that behaviours reported within the 12-month recall window occurred after circumcision for exposed individuals. Behavioural measures were therefore interpreted as post-circumcision outcomes within the reconstructed follow-up period.

For each outcome, analyses were restricted to men with valid responses to the relevant survey question. The proportion of true missing data was minimal across outcomes (≤0.7%) and did not differ materially by MMC status. Larger reductions in analytic sample size for some outcomes reflected outcome-specific eligibility criteria rather than missing data. For example, condom use at last sexual intercourse was defined only among men who reported having had sexual intercourse and who were asked the condom-use question in the survey. Outcome-specific denominators are therefore reported in the corresponding tables to distinguish eligibility restrictions from missing data.

#### Outcome measures: post-circumcision sexual behaviours

The primary outcomes were sexual behaviours hypothesised to occur after circumcision and potentially reflect behavioural adaptation following the procedure. These included condom use at last sexual intercourse, number of sexual partners in the past 12 months and self-reported symptoms of sexually transmitted infections in the past 12 months. The DHS STI indicator is based on self-reported symptoms during the preceding 12 months, including genital sores or ulcers and genital discharge. Because these measures rely on symptom recognition and self-report rather than laboratory confirmation, they may be affected by recall error, under-reporting, and misclassification, and therefore should be interpreted as proxies for symptomatic sexually transmitted infections rather than clinically confirmed diagnoses (25). Each outcome was analysed separately to capture distinct dimensions of sexual risk behaviour.

As men with less than one year of follow-up after circumcision were excluded, behaviours reported within the 12-month recall window occurred after circumcision for exposed individuals.

Because these behaviours occur after circumcision, they were not treated as confounders and were not included in exposure adjustment models. Instead, they were interpreted as post-exposure outcomes or behavioural pathways potentially linking circumcision to longer-term HIV risk.

For each behavioural outcome, analyses were restricted to men with valid responses to the relevant survey question. The proportion of true missing data was minimal across outcomes (≤0.7%) and did not differ materially by MMC status. The larger proportion of observations excluded from some outcome analyses reflects outcome-specific eligibility criteria rather than missingness. For example, the measure of condom use at last sexual intercourse was only defined among men who reported having had sexual intercourse and who were asked the condom-use question in the survey. Consequently, the effective analytic denominator for this outcome was smaller than the full cohort. Outcome-specific denominators used in each analysis are therefore reported explicitly in the corresponding tables to distinguish eligibility restrictions from missing data.

### Statistical data analysis

#### Conceptual framework and causal model

The statistical analysis was guided by an a priori causal framework formalised using a directed acyclic graph (DAG) (**Fig 2**). DAGs provide a transparent and principled approach for encoding identifying assumptions about causal structure and measured confounding, and for determining minimally sufficient adjustment sets in observational studies (20,26,27). However, as with all observational analyses, valid causal interpretation depends on the assumption that all relevant confounders are adequately measured and correctly specified; residual unmeasured confounding cannot be excluded (26–29).

**Fig 2.**
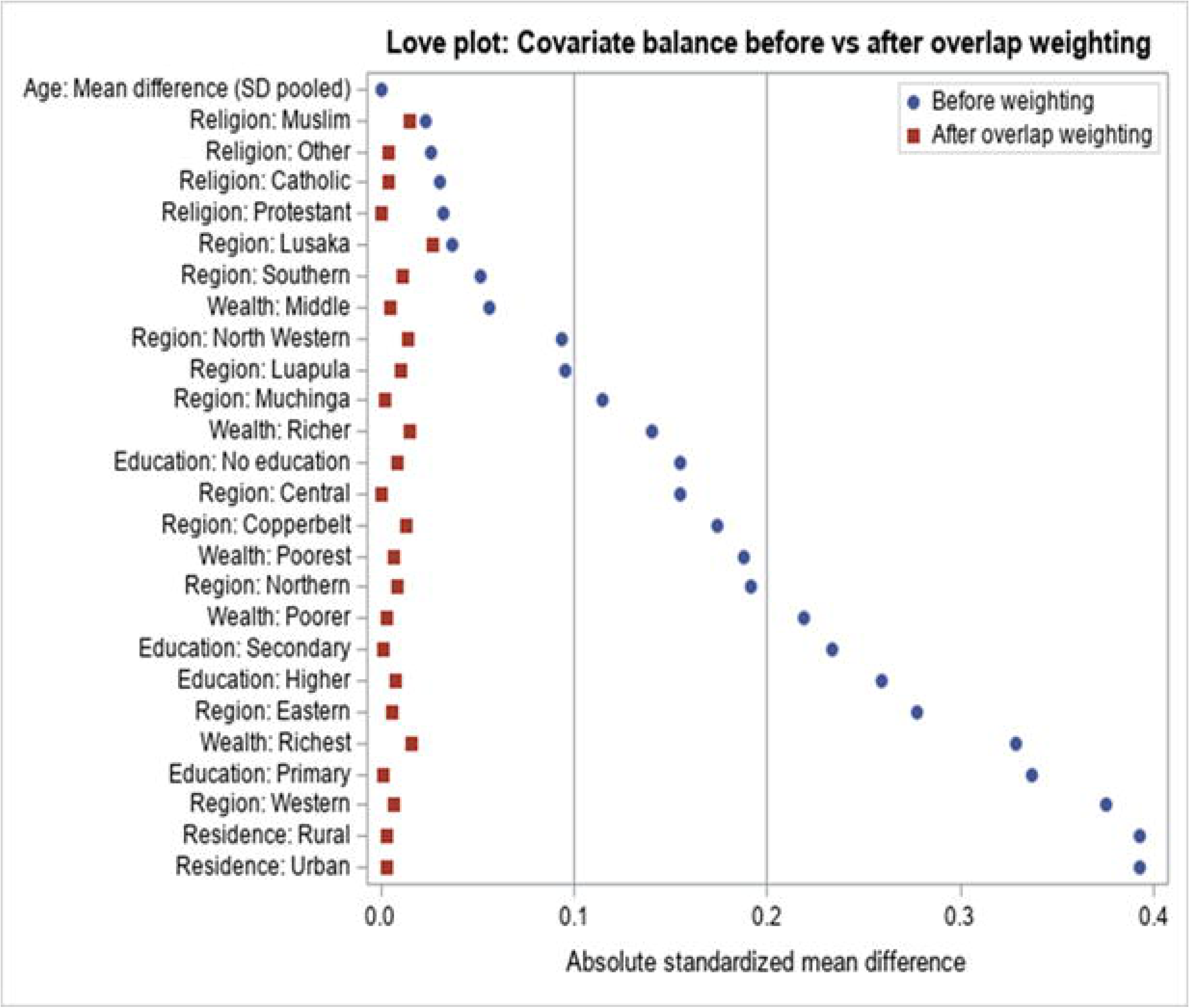
Directed acyclic graph illustrating the assumed causal structure between medical male circumcision and post-circumcision sexual behaviour outcomes. The diagram represents the conceptual framework used to guide covariate selection and causal analysis. Medical male circumcision (MMC) is specified as the exposure, and sexual behaviour outcomes—including condom non-use at last sexual intercourse, multiple sexual partners in the past 12 months, self-reported sexually transmitted infection (STI) in the past 12 months, and composite sexual risk behaviour indicators (SRBI ≥1 and SRBI ≥2)—are treated as post-exposure outcomes. Baseline characteristics, including age, region of residence, religion, place of residence (urban/rural), educational attainment, and household wealth quintile, are depicted as common causes of both circumcision uptake and sexual behaviour outcomes.

MMC was specified as the exposure of interest, and sexual behaviour outcomes—including condom non-use at last sex, multiple sexual partners in the past 12 months, self-reported sexually transmitted infection (STI) in the past 12 months, SRBI ≥1, and SRBI ≥2—were specified as binary outcomes.

Based on substantive knowledge and prior epidemiological evidence, age, region of residence, religion, place of residence (urban/rural), educational attainment, and household wealth quintile were identified as potential confounders. These variables were assumed to influence both the likelihood of undergoing MMC and subsequent sexual behaviour, while not lying on the causal pathway between MMC and the outcomes. The covariate set implied by the DAG was used consistently across all causal analyses.

#### Target trial emulation framework

The analytic design was informed by principles of target trial emulation (19). Conceptually, the emulated trial would include men aged 15–59 years participating in the ZDHS who reported their circumcision status and age at circumcision. The exposure strategies correspond to undergoing medical male circumcision at a given age versus remaining uncircumcised. Baseline was defined as the age at circumcision for circumcised men and the corresponding assigned baseline age for matched uncircumcised men, with follow-up extending from baseline to the age at survey interview. Outcomes comprised sexual behaviour indicators measured after baseline, including condom non-use at last sexual intercourse, multiple sexual partners in the previous 12 months, and self-reported STI symptoms. The causal contrast therefore compares these outcomes between circumcised and uncircumcised men with similar baseline characteristics, using overlap weighting to adjust for confounding. This framework clarifies the temporal ordering between exposure and behavioural outcomes in the observational data.

#### Descriptive analysis

Baseline age, follow-up time since baseline, and other baseline characteristics of the study population were summarised overall and by MMC status using both unweighted and survey-weighted estimates. Unweighted frequencies were determined to describe the observed sample composition, while survey-weighted estimates were used to characterise the target population represented in the study.

Continuous variables were summarised using weighted means with corresponding standard errors and 95% confidence intervals (CIs), accounting for the complex survey design, including stratification, clustering, and sampling weights. Medians and interquartile ranges were additionally reported using unweighted estimates to describe the distributional properties of key time-related variables. Categorical variables were summarised using weighted proportions with 95% CIs, estimated using survey-adjusted procedures. For categorical variables stratified by MMC status, proportions were calculated within MMC groups to facilitate comparison of distributions across exposure categories.

All survey-weighted descriptive analyses incorporated the DHS sampling weights and design variables to ensure valid population-level inference.

#### Propensity score estimation

Propensity scores were estimated to represent the conditional probability of undergoing MMC given observed baseline covariates. Specifically, the propensity score was defined as

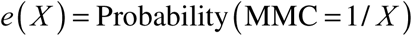

where *X* denotes the vector of baseline covariates. Logistic regression was used to model MMC status as a function of age, region of residence, religion, place of residence (urban/rural), educational attainment, and household wealth quintile. These variables were selected a priori based on substantive knowledge and the causal structure specified in the DAG (**Fig 3**).

**Fig 3.**
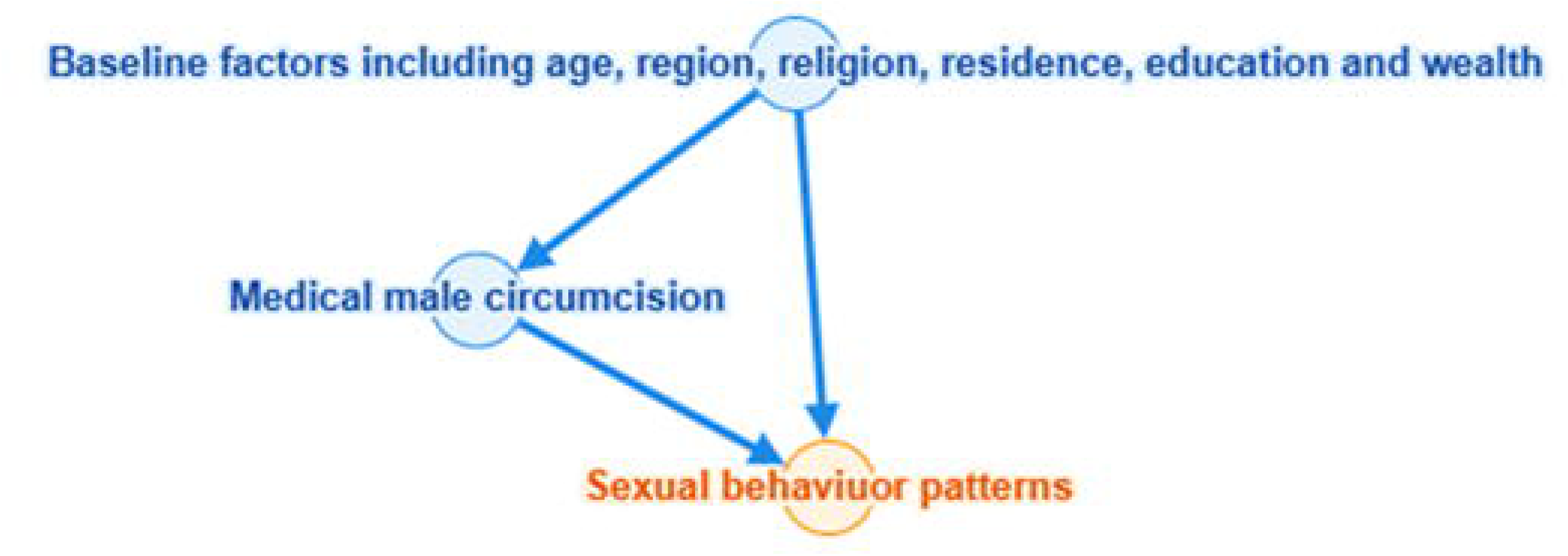
Distribution of estimated propensity scores by medical male circumcision status. The figure displays histograms and kernel density curves of estimated propensity scores for medically circumcised (treat = 1) and uncircumcised (treat = 0) men derived from the propensity score model. The distributions illustrate the degree of covariate overlap between exposure groups within the analytic sample and indicate the region of common support used to construct overlap weights for causal effect estimation.

Variables such as marital status, age at sexual debut, and HIV knowledge were not included because their timing relative to circumcision cannot be established in the cross-sectional data and they may occur after circumcision, potentially introducing post-treatment bias.

Consistent with established guidance for propensity score analyses, outcome variables were not included in the propensity score model in order to avoid model feedback and bias (22–24).

#### Overlap weighting

Overlap weighting was applied to address confounding and improve covariate balance between circumcised and uncircumcised men. For each individual *i* , overlap weights were defined as a function of the estimated propensity score *e_i_* as follows:

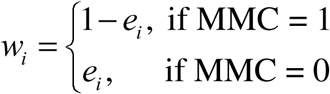

This weighting scheme assigns greater weight to individuals with substantial overlap in covariate distributions between exposure groups and down-weights individuals with extreme propensity scores. As a result, overlap weighting targets the average treatment effect in the overlap population and yields exact mean balance of included covariates in expectation when the propensity score model is correctly specified (30). The causal estimand therefore corresponds to the average treatment effect in the overlap population (ATO), representing men whose baseline characteristics are compatible with both circumcision and non-circumcision exposure states.

Overlap weighting was selected because it yields exact mean balance of included covariates in expectation and is less sensitive to extreme weights (30–32). To preserve population representativeness, the overlap weights were multiplied by the DHS sampling weights to generate a final analysis weight. Inference therefore pertains to the overlap population embedded within the national survey design, rather than the full target population. The combined weights were normalised, and variance estimation incorporated DHS stratification and clustering using Taylor series linearisation (33). All subsequent analyses incorporated this combined weight and accounted for survey stratification and clustering.

Inference from the weighted analyses therefore pertains to the *overlap population*—that is, men whose baseline characteristics are compatible with both circumcised and uncircumcised exposure groups—rather than to the entire male population of Zambia.

#### Covariate balance diagnostics

Covariate balance before and after weighting was assessed using standardized mean differences (SMDs). The SMD is a scale-invariant measure of covariate imbalance that is independent of sample size and is recommended for evaluating balance in propensity score analyses (24,34). Absolute SMD values below 0.10 were considered indicative of adequate balance.

Balance diagnostics were conducted for all covariates included in the propensity score model, including continuous variables and all levels of categorical variables, and were summarized using graphical inspection of overlap weight distributions and standardized SMD plots (Love plots).

#### Estimation of sexual behaviour prevalence

Weighted prevalence of sexual behaviour outcomes by MMC status was estimated using survey-adjusted cross-tabulations. These analyses incorporated the combined overlap and sampling weights and accounted for stratification and clustering. Outcome-specific analyses were conducted using complete-case data, with exclusions due to missingness handled consistently across outcomes.

#### Effect estimation

Associations between MMC and sexual behaviour outcomes were estimated using logistic regression models incorporating overlap weights and accounting for the complex survey design. The primary estimands were odds ratios (ORs) comparing circumcised and uncircumcised men within the overlap population defined by the weighting scheme.

Two model specifications were considered. First, marginal models including MMC as the sole predictor were fitted to estimate weighted population-average causal effects. Second, covariate-adjusted models were fitted that included age, region, religion, residence, education, and wealth quintile to improve precision and assess robustness of the MMC effect. In all models, MMC was coded as a binary exposure with uncircumcised men as the reference group.

#### Sensitivity analysis for unmeasured confounding

To assess the potential influence of unmeasured confounding on the estimated associations between MMC and behavioural outcomes, sensitivity analyses were conducted using evidence (E) -values. E-values quantify the minimum strength of association that an unmeasured confounder would need to have with both the exposure (MMC) and the outcome, conditional on the measured covariates, to fully explain away the observed association (35).

For each outcome, E-values were calculated based on the adjusted odds ratios obtained from the overlap-weighted logistic regression models. When the point estimate indicated a protective association (odds ratio <1), the reciprocal of the odds ratio was used in accordance with established guidance for computing E-values for protective effects (36). Larger E-values indicate that stronger associations of an unmeasured confounder with both the exposure and the outcome would be required to explain away the observed association. Conversely, E-values close to 1 indicate that relatively modest confounding associations could potentially account for the estimate. Interpretation of E-values therefore focused on the magnitude of confounding that would be required to explain the observed association.

#### Statistical software and inference

All analyses were conducted using SAS software (version 9.4). Survey-specific procedures were used throughout to appropriately account for the complex sampling design. Statistical significance was assessed using two-sided tests at the 5% level.

#### Ethical considerations

The DHS data are publicly available and fully anonymised prior to release. Ethical approval for the original survey was obtained by the DHS Program from relevant national and institutional review boards. The present study involved secondary analysis of de-identified data and its ethical approval was obtained from the National Health Research and Training Institute (NHRTI) Ethics Review Committee (ERC) (NHRTREC/23/03/26).

#### Use of artificial intelligence tools and technologies

Artificial intelligence tools (ChatGPT, OpenAI) were used solely to assist with language editing using the prompt, “edit sentences.” They were not used in study design, data analysis, or generation of results. All AI-generated outputs were reviewed and validated by the authors, who take full responsibility for the accuracy, originality, and integrity of the manuscript.

## Results

### Study population and baseline characteristics

The resulting matched pseudo-cohort comprised 9,609 men, of whom 6,406 (66.7%) were uncircumcised and 3203 (33.3%) were medically circumcised (**Table 1**). Because uncircumcised men were matched to circumcised men on current age and assigned a common baseline age anchored to the age at circumcision, baseline age was effectively aligned across groups. Accordingly, the mean baseline age was 16.5 years in both groups, with identical medians (15.0 years) and interquartile ranges (12 to 19 years), and not statistically significantly different (p = 0.460). Overall, the mean follow-up time was 11.2 years; the mean was 11.2 years among uncircumcised men and 11.1 years among circumcised men, with no statistically significant difference between the groups (p = 0.926). Across follow-up time categories, the largest proportion of observations fell within the 5–9 year category (31.3% [95% CI: 30.3–32.3] overall), comprising 66.1% (95% CI: 63.8–68.3) of uncircumcised men and 33.9% (95% CI: 31.7–36.2) of circumcised men; the distribution of follow-up time categories did not differ significantly between groups (p = 0.663).

**Table 1.**
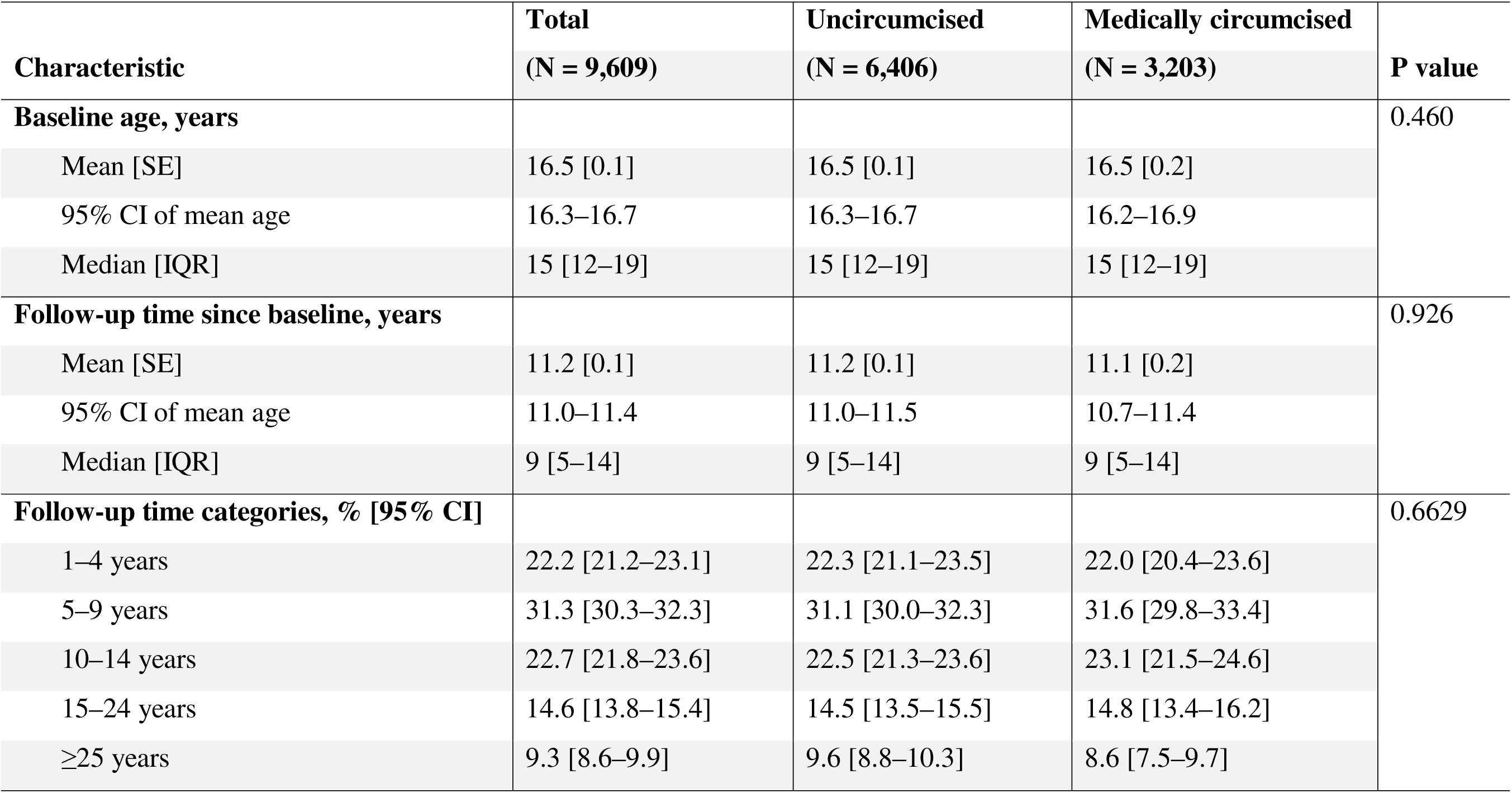
Baseline age and follow-up time characteristics of the matched pseudo-cohort by medical male circumcision status.

The overall mean age was 27.7 years (**Table 2**). Before overlap-weighting, circumcised men were slightly younger than uncircumcised men (mean age 27.6 vs 27.7 years). After overlap weighting, mean age was similar between groups (27.8 years among circumcised men and 28.0 years among uncircumcised men). In the unweighted data, marked differences in sociodemographic characteristics were observed by MMC status. Circumcised men were more likely to reside in urban areas (51.7% vs 32.7%), to have attained secondary or higher education (69.0% vs 50.1%), and to belong to the richest wealth quintile (26.0% vs 13.2%). Regional distributions also differed substantially, with circumcised men overrepresented in Lusaka (11.7% vs 10.6%) and Western provinces (15.6% vs 4.5%) and underrepresented in Eastern (6.4% vs 14.8%) and Northern provinces (5.5% vs 10.7%).

**Table 2.**
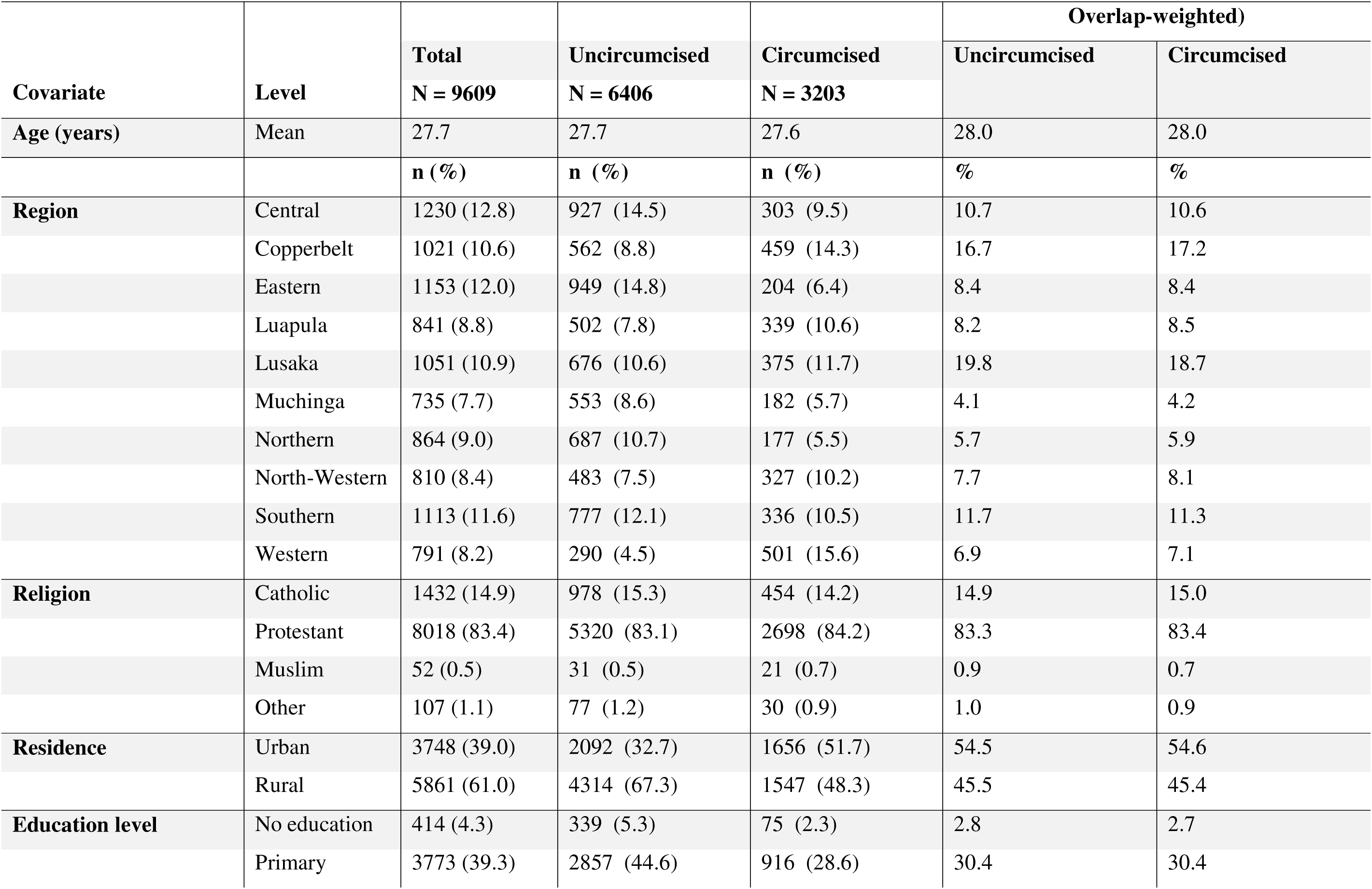
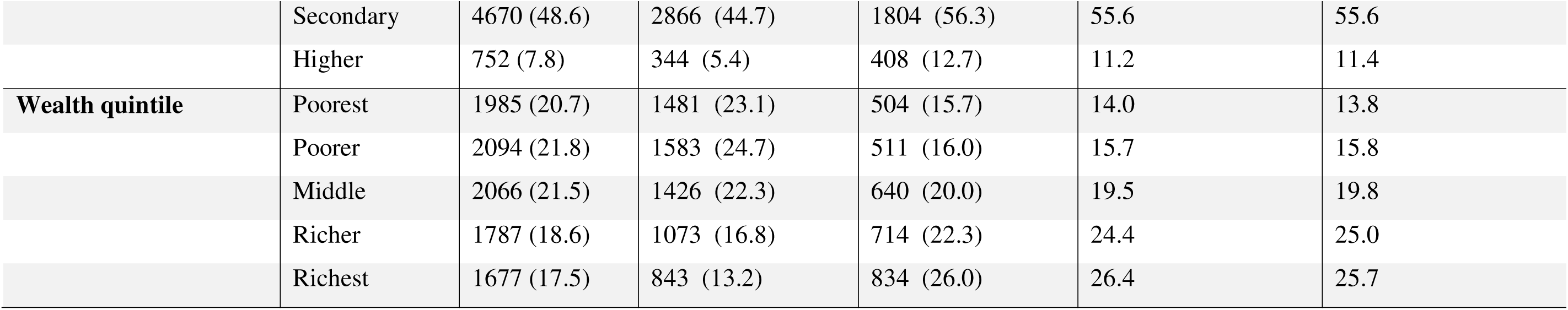
Baseline characteristics of the study population by medical male circumcision status, before and after overlap weighting.

After applying overlap weights, the distributions of all baseline covariates were closely aligned between circumcised and uncircumcised men. For example, the overlap-weighted proportion residing in urban areas was 54.6% among circumcised men and 54.5% among uncircumcised men, while the overlap-weighted proportion in the richest wealth quintile was 25.7% and 26.4%, respectively. Similar alignment was observed across region, religion, education level, and wealth quintile, indicating improved covariate balance after weighting.

### Prevalence of sexual behaviour outcomes

**Table 3** presents overlap-weighted prevalence estimates for sexual behaviour outcomes by MMC status within the analytic population defined by the overlap weighting scheme. In the unweighted sample, condom non-use was reported by 74.6% of uncircumcised men and 66.9% of circumcised men. After applying overlap weighting, the prevalence remained higher among uncircumcised men (72.0%) than circumcised men (66.9%).

**Table 3.**
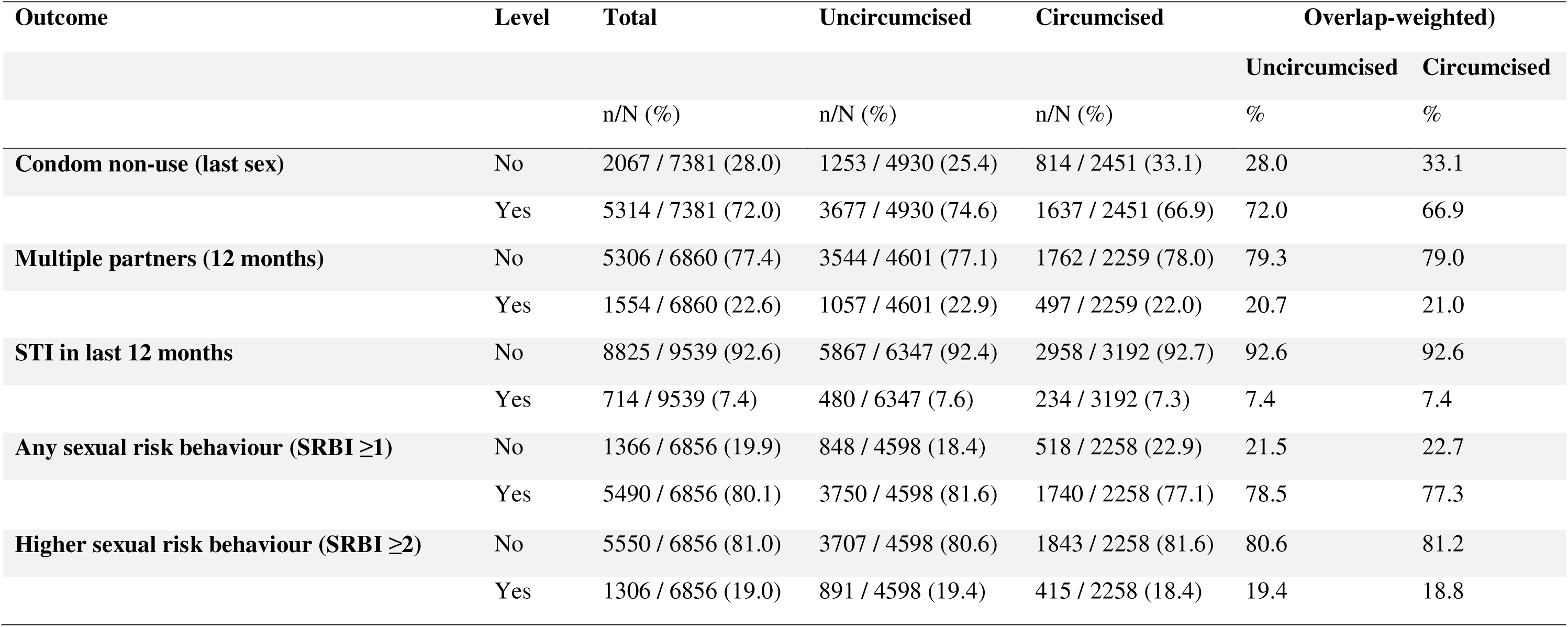
Prevalence of sexual behaviour outcomes by medical male circumcision status, unweighted counts and overlap-weighted percentages.

Reporting multiple sexual partners in the previous 12 months was less common. In the unweighted sample, 22.9% of uncircumcised men and 22.0% of circumcised men reported having two or more partners. After overlap weighting, the estimated prevalence was 20.7% among uncircumcised men and 21.0% among circumcised men.

Self-reported STI symptoms in the previous 12 months were relatively uncommon. In the analytic sample, 714 of 9,539 men reported STI symptoms, corresponding to a prevalence of 7.4%. Overlap-weighted estimates were similar between groups, with a prevalence of 7.4% among both circumcised and uncircumcised men.

SRBI ≥ 1 was frequently reported. In the unweighted sample, 81.6% of uncircumcised men and 77.1% of circumcised men reported at least one sexual risk behaviour. After overlap weighting, the prevalence of SRBI ≥ 1 was 78.5% among uncircumcised men and 77.3% among circumcised men.

SRBI ≥ 2 was less common. The overlap-weighted prevalence was 18.8% among circumcised men and 19.4% among uncircumcised men.

### Overlap-weighted associations between MMC and sexual behaviours

**Table 4** summarises overlap-weighted effect estimates from survey-adjusted logistic regression models. Within the overlap population defined by the weighting scheme, MMC was associated with lower odds of condom non-use at last sex, with an OR of 0.785 (95% CI: 0.699–0.883, p < 0.0001). No association was observed between MMC and reporting multiple sexual partners in the past 12 months (OR = 1.020, 95% CI: 0.887–1.174, p = 0.779). MMC was also not associated with self-reported STI in the past 12 months (OR = 1.042, 95% CI: 0.868–1.251, p = 0.659). MMC was associated with reduced odds of SRBI ≥ 1 (OR = 0.845, 95% CI: 0.735–0.972, p = 0.018). No statistically significant association was observed for SRBI ≥ 2.

**Table 4.**
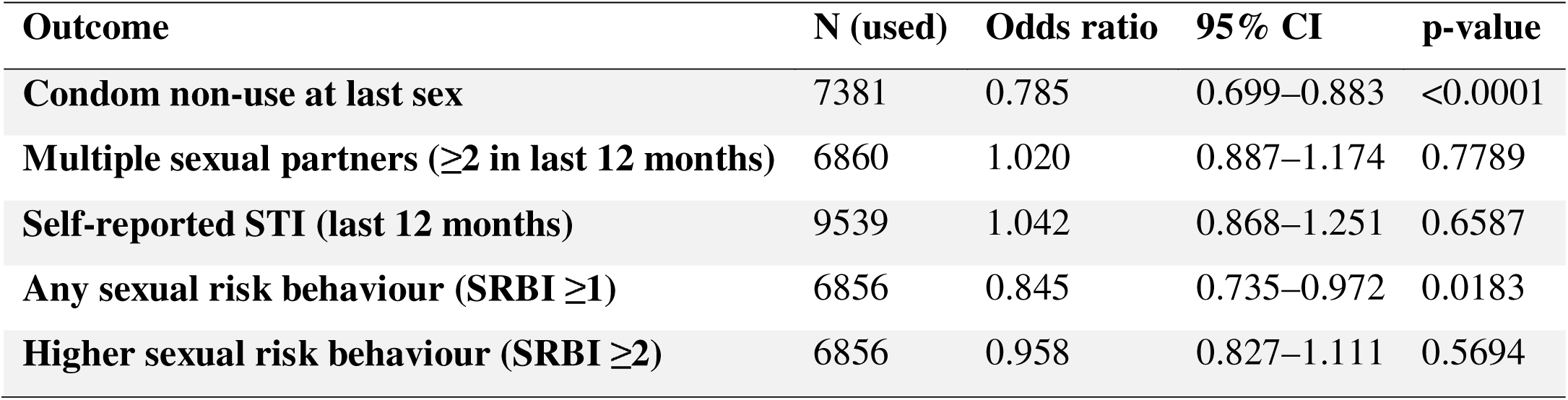
Overlap-weighted survey-adjusted effects of medical male circumcision on sexual behaviours.

**Table 5** presents overlap-weighted survey-adjusted outcome models that additionally adjusted for age, region, religion, residence, education level, and wealth quintile. After covariate adjustment, MMC remained associated with lower odds of condom non-use at last sex (adjusted OR [aOR] = 0.753, 95% CI: 0.668–0.850, p < 0.001). No statistically significant associations were observed between MMC and multiple sexual partners (aOR = 1.004, 95% CI: 0.873–1.156, p = 0.953) or self-reported STI (aOR = 1.046, 95% CI: 0.868–1.259, p = 0.637). The inverse association between MMC and SRBI ≥ 1 persisted after covariate adjustment (aOR = 0.825, 95% CI: 0.715–0.951, p = 0.008). MMC was not significantly associated with SRBI ≥ 2 in adjusted models (aOR = 0.937, 95% CI: 0.808–1.086, p = 0.388).

**Table 5.** Overlap-weighted survey-adjusted effects of male medical circumcision on sexual behaviours from outcome models adjusted for covariates.

| **Outcome** | **N (used)** | **Adjusted Odds Ratio** | **95% CI** | **p-value** |
| --- | --- | --- | --- | --- |
| **Condom non-use at last sex** | 7381 | 0.753 | 0.668–0.850 | <0.0001 |
| **Multiple partners (last 12 months; $\geq 2$)** | 6860 | 1.004 | 0.873–1.156 | 0.9529 |
| **Self-reported STI (last 12 months)** | 9539 | 1.046 | 0.868–1.259 | 0.6368 |
| **Any sexual risk behaviour (SRBI $\geq 1$)** | 6856 | 0.825 | 0.715–0.951 | 0.0080 |
| **Higher sexual risk behaviour (SRBI $\geq 2$)** | 6856 | 0.937 | 0.808–1.086 | 0.3881 |

### Diagnostic assessment of overlap weighting

**Fig 3** shows the distribution of estimated propensity scores by medical male circumcision status. Prior to weighting, the propensity score distributions for circumcised and uncircumcised men differed in their central tendency, with circumcised men having higher estimated probabilities of circumcision. Despite these differences, the distributions overlapped across a wide range of propensity score values, particularly within the central region of the score distribution. This region of overlap represents the area of common support, where individuals with similar baseline characteristics exist in both circumcised and uncircumcised groups and therefore contribute most strongly to the overlap-weighted causal effect estimates.

Standardised mean differences for baseline covariates before and after application of overlap weights are presented in **Table 6**. Before weighting, several covariates displayed substantial imbalance between circumcised and uncircumcised men. Absolute standardised mean differences exceeded 0.20 for multiple categories of education, region, wealth quintile, and place of residence. The largest unweighted imbalances were observed for urban residence (SMD = 0.393), Western Province (SMD = 0.376), richest wealth quintile (SMD = 0.329), and primary education level (SMD = −0.337).

**Table 6.** Standardised mean differences before and after overlap weighting.

| Covariate | Level | SMD (Unweighted) | SMD (Overlap-weighted) |
| --- | --- | --- | --- |
| Education | Higher | 0.259 | 0.008 |
|  | No education | −0.154 | −0.008 |
|  | Primary | −0.337 | −0.001 |
|  | Secondary | 0.233 | −0.001 |
| **Region** | Central | −0.155 | 0.001 |
|  | Copperbelt | 0.175 | 0.013 |
|  | Eastern | −0.277 | −0.005 |
|  | Luapula | 0.095 | 0.010 |
|  | Lusaka | 0.037 | −0.027 |
|  | Muchinga | −0.115 | 0.002 |
|  | North-Western | 0.094 | 0.015 |
|  | Northern | −0.191 | 0.009 |
|  | Southern | −0.052 | −0.012 |
|  | Western | 0.376 | 0.006 |
| **Religion** | Catholic | −0.031 | 0.004 |
|  | Muslim | 0.023 | −0.015 |
|  | Other | −0.026 | −0.003 |
|  | Protestant | 0.032 | 0.000 |
| **Residence** | Rural | −0.393 | −0.003 |
|  | Urban | 0.393 | 0.003 |
| **Wealth quintile** | Middle | −0.056 | 0.005 |
|  | Poorer | −0.219 | 0.003 |
|  | Poorest | −0.187 | −0.007 |
|  | Richer | 0.140 | 0.015 |
|  | Richest | 0.329 | −0.016 |

After application of overlap weights, standardised mean differences were markedly reduced across all covariates. For categorical covariates, absolute SMDs ranged from 0.000 to 0.027 following weighting. For example, the SMD for urban residence decreased from 0.393 before weighting to 0.003 after weighting, for Western Province from 0.376 to 0.006, and for the richest wealth quintile from 0.329 to −0.016.

**Fig 4** summarises covariate balance before and after overlap weighting using absolute standardised mean differences. Prior to weighting, several covariates exceeded the conventional imbalance threshold of 0.10. After application of overlap weights, all covariates were reduced to absolute standardized mean differences below the conventional threshold of 0.10, indicating substantial reduction in baseline imbalance between exposure groups.

**Fig 4.**
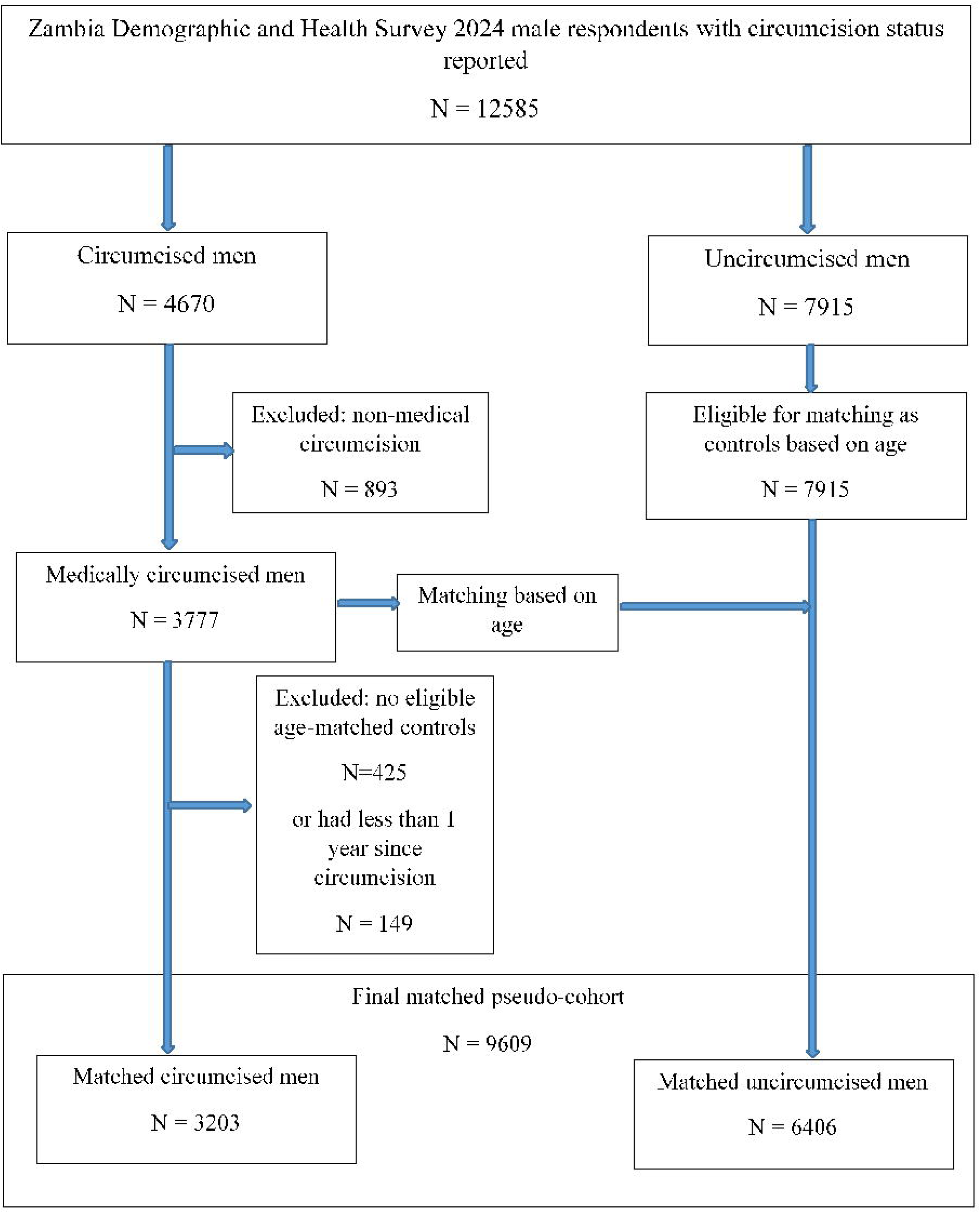
Covariate balance before and after application of overlap weighting. Love plot showing absolute standardised mean differences (SMDs) for baseline covariates before weighting and after application of overlap weights derived from the propensity score model. Points represent the magnitude of imbalance for each covariate and category included in the propensity score model. The horizontal reference line indicates the conventional threshold for acceptable covariate balance (|SMD| = 0.10).

### Sensitivity to unmeasured confounding

**Table 7** presents the results of the sensitivity analysis assessing the potential influence of unmeasured confounding on the association between MMC and behavioural outcomes. The E-value for the association between MMC and condom non-use at last sex was 1.99, indicating that an unmeasured confounder would need to be associated with both circumcision status and condom use by a risk ratio of approximately two-fold to fully explain away the observed association. Similarly, the E-value for the association between MMC and any sexual risk behaviour (SRBI ≥1) was 1.72, suggesting moderate robustness to unmeasured confounding.

**Table 7.** Sensitivity analysis using E-values for potential unmeasured confounding.

| **Outcome** | **Adjusted Odds Ratio** | **95% CI** | **E-value** | **Interpretation** |
| --- | --- | --- | --- | --- |
| **Condom non-use at last sex** | 0.753 | 0.668–0.850 | 1.99 | Moderate robustness |
| **Multiple sexual partners ($\geq 2$ in last 12 months)** | 1.004 | 0.873–1.156 | 1.07 | Null association |
| **Any sexual risk behaviour (SRBI $\geq 1$)** | 0.825 | 0.715–0.951 | 1.72 | Moderate robustness |
| **Higher sexual risk behaviour (SRBI $\geq 2$)** | 0.937 | 0.808–1.086 | 1.33 | Weak robustness |

In contrast, the E-values for multiple sexual partners (1.07) and higher sexual risk behaviour (SRBI ≥2; 1.33) were small, reflecting the null or weak associations observed for these outcomes.

## Discussion

Using nationally representative data from ZDHS, this study examined the association between MMC and subsequent sexual behaviours, while demonstrating a causal analytic approach for evaluating behavioural outcomes using cross-sectional survey data. By reconstructing temporal ordering through age at circumcision and applying overlap weighting within a matched pseudo-cohort framework, the analysis addresses a central methodological challenge in observational evaluations of MMC: the misclassification of post-circumcision sexual behaviours as baseline confounders.

The pseudo-cohort framework relies on several assumptions common to causal analyses using observational data. First, the design assumes that uncircumcised men matched to circumcised men were eligible to undergo circumcision at the assigned baseline age, thereby representing the counterfactual exposure state. Second, the validity of causal interpretation depends on adequate measurement of confounders influencing both circumcision uptake and sexual behaviour. Finally, because the reconstruction is based on age rather than calendar time, behavioural outcomes could potentially vary across birth cohorts. Although exact matching on age at interview reduces these differences, residual cohort effects cannot be entirely excluded.

As designed, the matched pseudo-cohort achieved close comparability between circumcised and uncircumcised men with respect to baseline age and accumulated follow-up time. The similarity of both continuous measures and follow-up time distributions indicates that men in the two groups were observed over comparable post-baseline periods. This temporal alignment is important for behavioural analyses, as it limits confounding arising from differences in age structure or time since exposure and provides a coherent foundation for interpreting subsequent behavioural comparisons (20,21,37).

### Behavioural outcomes under routine programme conditions

Overall, the findings did not indicate increased sexual risk behaviour among circumcised men within the overlap population analysed in this study. Instead, patterns of sexual behaviour among circumcised men were broadly similar to, and in some cases more protective than, those observed among uncircumcised men. In particular, circumcised men did not report higher levels of multiple sexual partnerships or sexually transmitted infection symptoms, and composite measures of sexual risk were not elevated. These findings provide no support for behavioural risk compensation following MMC and are consistent with the behavioural neutrality observed in earlier randomised and prospective cohort studies, while extending that evidence to a contemporary, population-based programme setting.

Substantively, these findings are consistent with the behavioural conclusions of the landmark RCTs conducted in South Africa, Kenya, and Uganda, which demonstrated substantial biological protection against HIV acquisition without corresponding increases in sexual risk behaviour among circumcised men (38–40). Long-term follow-up of these trial cohorts similarly found no evidence of risk compensation over time (39,41). However, the relevance of these trial-era findings to contemporary, large-scale circumcision programmes implemented under routine health-system conditions has remained an open question, particularly as HIV prevention landscapes have evolved.

The lower odds of condom non-use observed among circumcised men in this analysis may plausibly reflect the structure of modern VMMC programmes, which incorporate HIV prevention counselling and risk-reduction messaging as routine components of service delivery. Several programme evaluations and cohort studies from eastern and southern Africa have reported stable or improved condom use among circumcised men following VMMC, suggesting that counselling delivered during circumcision services may reinforce safer sexual practices rather than promote behavioural risk compensation (5,6,42). The present findings suggest that these programme-level effects are observable at the population level in Zambia.

In contrast, the absence of associations between MMC and multiple sexual partnerships or STI symptoms aligns with recent population-based analyses using DHS and cohort data across sub-Saharan Africa, which generally report behavioural neutrality once sociodemographic differences are appropriately addressed (43). While self-reported STI measures are inherently limited by recall and reporting biases, the null association observed here is reassuring in the context of persistent concerns that MMC might encourage higher-risk partner acquisition.

The methodological contribution of this study lies in demonstrating why observational analyses have often struggled to reproduce trial findings and how this limitation can be addressed using existing population survey data. Much of the post-trial observational literature has relied on conventional regression adjustment strategies that do not explicitly account for the temporal ordering of exposure and behaviour, thereby risking the treatment of behaviours measured at the time of survey as pre-exposure confounders (19,20). As highlighted in recent causal inference scholarship, adjusting for variables that occur after exposure can introduce post-treatment bias, distort effect estimates, and obscure true intervention effects (15,20).

By explicitly reconstructing temporality through age at circumcision and treating sexual behaviours as post-exposure outcomes rather than baseline confounders, this study aligns the analytic structure of DHS data with the underlying causal process of MMC delivery and behavioural response. The application of overlap weighting further strengthens causal interpretation by focusing inference on men with comparable baseline characteristics and avoiding the instability associated with extreme inverse probability weights (30,31). Notably, when these modern causal tools are applied, the resulting observational estimates closely converge with the conclusions of randomised and prospective cohort studies.

This convergence is particularly important given that contemporary circumcision programmes operate in markedly different epidemiological and social contexts from early efficacy trials, including widespread antiretroviral therapy coverage, expanded HIV testing, and evolving norms around masculinity and sexual risk (44–47). Recent cohort and modelling studies have emphasised that behavioural responses to prevention interventions are context-dependent and may change over time (48–50). The present findings suggest that, even within these changing contexts, behavioural risk compensation following MMC is not evident when analyses respect temporal ordering.

### Implications for research and surveillance

For research, the study highlights the value of explicitly addressing temporality and post-exposure bias when analysing behavioural outcomes in cross-sectional surveys. The analytic framework demonstrated here is applicable beyond MMC—for example, in evaluations of HIV testing, pre-exposure prophylaxis uptake, or other preventive interventions where age at first exposure is recorded. Future work could extend this approach using Bayesian evidence-synthesis methods or by combining DHS data with routine health information systems to further strengthen causal inference and improve the precision of population-level estimates.

More broadly, these findings support the use of DHS data as an active component of cumulative causal evidence rather than as a purely descriptive or confirmatory data source. In settings where randomised trials are no longer ethical, feasible, or timely, robust observational methods are essential for monitoring long-term behavioural and programmatic effects of HIV prevention interventions.

### Strengths and limitations

This study has several strengths. It uses a large, nationally representative sample and applies an explicit causal framework informed by directed acyclic graphs to guide covariate selection and analysis. The study also incorporates rigorous covariate balance diagnostics and fully accounts for the complex survey design of the Demographic and Health Survey. The consistency of findings across marginal and covariate-adjusted overlap-weighted models supports the stability of the estimates. In addition, sensitivity analyses using E-values indicated that moderately strong unmeasured confounding would be required to fully explain the observed associations for key outcomes.

Several limitations should also be considered. First, the matched pseudo-cohort required exclusion of circumcised men without eligible age-matched uncircumcised controls, which may introduce selection bias if excluded men differed systematically from those retained. As shown in S1 Table, excluded men were younger and circumcised more recently, resulting in shorter follow-up time. Consequently, the findings primarily reflect behavioural patterns among men with at least one year of follow-up after circumcision and may be less generalisable to very recently circumcised cohorts. Second, despite the pseudo-cohort framework, residual unmeasured confounding cannot be excluded. Individual-level factors such as risk preferences or partner characteristics are not fully captured in DHS data. Third, circumcision status, age at circumcision, and sexual behaviour outcomes were self-reported and may therefore be subject to recall error and social desirability bias. Misclassification of circumcision status or differential reporting of sexual behaviour could occur, particularly in settings where both traditional and medical circumcision are practiced. In addition, counselling delivered during voluntary medical male circumcision services emphasises risk-reduction behaviours, which may influence how circumcised men report sexual practices in surveys. Finally, the estimated effects apply to the overlap population and may not generalise to men with very high or very low propensity for circumcision.

Several features of the analysis help mitigate these concerns. Diagnostic comparisons indicated that circumcised men excluded during pseudo-cohort construction differed primarily in age and follow-up time, while the distributions of region, residence, religion, and household wealth were broadly similar between excluded and retained individuals, suggesting limited socioeconomic selection. Overlap weighting achieved excellent covariate balance across all measured baseline characteristics, with absolute standardised mean differences reduced to below conventional thresholds, indicating that circumcised and uncircumcised men were highly comparable within the analytic population. The consistency of findings across marginal and covariate-adjusted models, together with the E-value sensitivity analyses, suggests that relatively strong unmeasured confounding would be required to fully explain the observed association between circumcision and condom use. Finally, the large nationally representative DHS sample and the explicit reconstruction of temporality using age at circumcision strengthen internal validity relative to conventional cross-sectional analyses that treat behavioural variables as baseline confounders.

## Conclusion

This study found no evidence of increased sexual risk behaviour among circumcised men compared with uncircumcised men within the overlap population. On the contrary, MMC was associated with lower odds of condom non-use and any reported sexual risk behaviour among men in the overlap population represented in the matched pseudo-cohort, while showing no association with multiple partnerships or STI symptoms. Beyond these substantive findings, the study demonstrates that cross-sectional population surveys—when analysed using causal frameworks that respect temporality and avoid post-treatment bias—can closely approximate benchmark trial conclusions. This methodological insight strengthens confidence in the use of DHS data for contemporary HIV prevention evaluation and supports the continued integration of MMC within combination prevention strategies.

## Data Availability

All data produced are available online at the Demographic Health Surveys program website.

https://www.dhsprogram.com/methodology/survey/survey-display-610.cfm

## Acknowledgements

We thank Professor Seter Siziya for his comprehensive comments on the initial draft, Lucy Ngulube Phiri and Bridget Musonda for their roles in data curation and secure storage, and the Integrated Public Use Microdata Series (IPUMS) and DHS Program for providing access to the data used in this study.

## Supporting information

**S1 Table. Comparison of medically circumcised men included and excluded from the matched pseudo-cohort.** Differences between included and excluded circumcised men were primarily driven by age and time since circumcision, reflecting the matching requirements of the pseudo-cohort design; younger and more recently circumcised men were more likely to be excluded, which also explains differences in educational attainment, while distributions of province, residence, religion, and wealth were similar, indicating no substantial socioeconomic or geographic selection bias. Overall, these findings suggest that observed differences largely reflect the age-dependent structure of the matching procedure rather than systematic socioeconomic selection.

