## Supplementary material for "Sexual risk behaviours following medical male circumcision: a matched pseudo-cohort analysis using population-based survey data": S1 Table

**Table S1. Comparison of medically circumcised men included and excluded from the matched pseudo-cohort**

| Characteristic | Excluded | Included | P-value |
| --- | --- | --- | --- |
|  | **(n = 425)** | **(n = 3203)** |  |
| Current age at interview, mean (SD) | 20.9 (3.41) | 27.5 (10.46) | <0.001 |
| Age at circumcision, mean (SD) | 14.5 (4.39) | 16.4 (7.86) | 0.003 |
| Potential follow-up time (years), mean (SD) | 6.44 (4.43) | 11.10 (9.21) | <0.001 |
| Region, n (%) |  |  | 0.534 |
| Central | 45 (10.6) | 303 (9.5) |  |
| Copperbelt | 64 (15.1) | 459 (14.3) |  |
| Eastern | 38 (8.9) | 204 (6.4) |  |
| Luapula | 38 (8.9) | 339 (10.6) |  |
| Lusaka | 53 (12.5) | 375 (11.7) |  |
| Muchinga | 18 (4.2) | 182 (5.7) |  |
| Northern | 25 (5.9) | 177 (5.5) |  |
| North-Western | 37 (8.7) | 327 (10.2) |  |
| Southern | 43 (10.1) | 336 (10.5) |  |
| Western | 64 (15.1) | 501 (15.6) |  |
| Residence, n (%) |  |  | 0.875 |
| Urban | 218 (51.3) | 1656 (51.7) |  |
| Rural | 207 (48.7) | 1547 (48.3) |  |
| Education level, n (%) |  |  | <0.001 |
| No education | 3 (0.7) | 75 (2.3) |  |
| Primary | 117 (27.5) | 916 (28.6) |  |
| Secondary | 284 (66.8) | 1804 (56.3) |  |
| Higher | 21 (4.9) | 408 (12.7) |  |
| Religion, n (%) |  |  | 0.601 |
| Catholic | 55 (12.9) | 454 (14.2) |  |
| Protestant | 364 (85.6) | 2698 (84.2) |  |
| Muslim | 4 (0.9) | 21 (0.7) |  |
| Other | 2 (0.5) | 30 (0.9) |  |
| Wealth quintile, n (%) |  |  | 0.878 |
| Poorest | 74 (17.4) | 504 (15.7) |  |
| Poorer | 65 (15.3) | 511 (16.0) |  |
| Middle | 83 (19.5) | 640 (20.0) |  |
| Richer | 89 (20.9) | 714 (22.3) |  |
| Richest | 114 (26.8) | 834 (26.0) |  |
